# Glaucoma Detection and Feature Visualization from OCT Images Using Deep Learning

**DOI:** 10.1101/2023.03.06.23286879

**Authors:** Nahida Akter, Stuart Perry, John Fletcher, Matthew P. Simunovic, Fiona Stapleton, Maitreyee Roy

## Abstract

**Purpose:** In this paper, we aimed to clinically interpret Temporal-Superior-Nasal-Inferior-Temporal (TSNIT) retinal optical coherence tomography (OCT) images in a convolutional neural network (CNN) model to differentiate between normal and glaucomatous optic neuropathy.

**Methods:** Three modified pre-trained deep learning (DL) models: SqueezeNet, ResNet18, and VGG16, were fine-tuned for transfer learning to visualize CNN features and detect glaucoma using 780 segmented and 780 raw TSNIT OCT B-scans of 370 glaucomatous and 410 normal images. The performance of the DL models was further investigated with Grad-CAM activation function to visualize which regions of the images are considered for the prediction of the two classes.

**Results:** For glaucoma detection, VGG16 performed better than SqueezeNet and ResNet18 models, with the highest AUC (0.988) on validation data and accuracy of 93% for test data. Moreover, identical classification results were obtained from raw and segmented images. For feature localization, three models accurately identify the distinct retinal regions of the TSNIT images for glaucoma and normal eyes.

**Conclusion:** This evidence-based result demonstrates the remarkable effectiveness of using raw TSNIT OCT B-scan for automated glaucoma detection using DL techniques which mitigates the black box problem of artificial intelligence (AI) and increases the transparency and reliability of the DL model for clinical interpretation. Moreover, the results imply that the raw TSNIT OCT scan can be used to detect glaucoma without any prior segmentation or pre-processing, which may be an attractive feature in large-scale screening applications.

## 1. Introduction

Glaucoma is a progressive optic neuropathy characterized by retinal ganglion cell (RGC) death, leading to neuroretinal rim tissue loss at the optic nerve and corresponding irreversible vision loss.^1^ It is one of the leading causes of blindness: 3.4% of people aged 40-80 years are estimated to suffer from glaucoma, and the global burden has been predicted to rise to around 112 million by the year 2040.^2^ Approximately 2.7 million people in the US were affected in 2010, and this is expected to increase by more than two-fold by 2050 (6.3 million).^3^ Early diagnosis is essential because promptly addressing modifiable risk factors, such as intraocular pressure and precipitating factors for its elevation (e.g., angle-closure), help to minimize vision loss. However, as few as 50% of all cases are diagnosed correctly in those who have seen an eye care practitioner in the past year.^4^

There is potential for AI to support glaucoma detection. However, there are particular challenges, especially in identifying differences in the optic nerve and retina of normal eyes from those with early glaucoma. It is assumed that most glaucomatous pathology is evident at the optic nerve head (ONH) and the peripapillary RGC layer due to characteristic axonal degeneration.^5^ Regardless of the underlying aetiological mechanisms at play, this damage results in so-called “peripapillary atrophy”, with thinning of the chorioretinal and retinal pigment epithelium (RPE) surrounding the optic nerve.^6^ OCT is a popular imaging technology that can generate a cross-sectional B-scan of the circumpapillary retinal regions from the centre of the ONH (Figure 1(a)). It facilitates monitoring the thickness variations of the retinal layer commencing at the temporal quadrant in a clockwise direction, referred to as the “temporal-superior-nasal-inferior-temporal” (TSNIT) retinal layers (Figure 1(b)).

**Figure 1.**
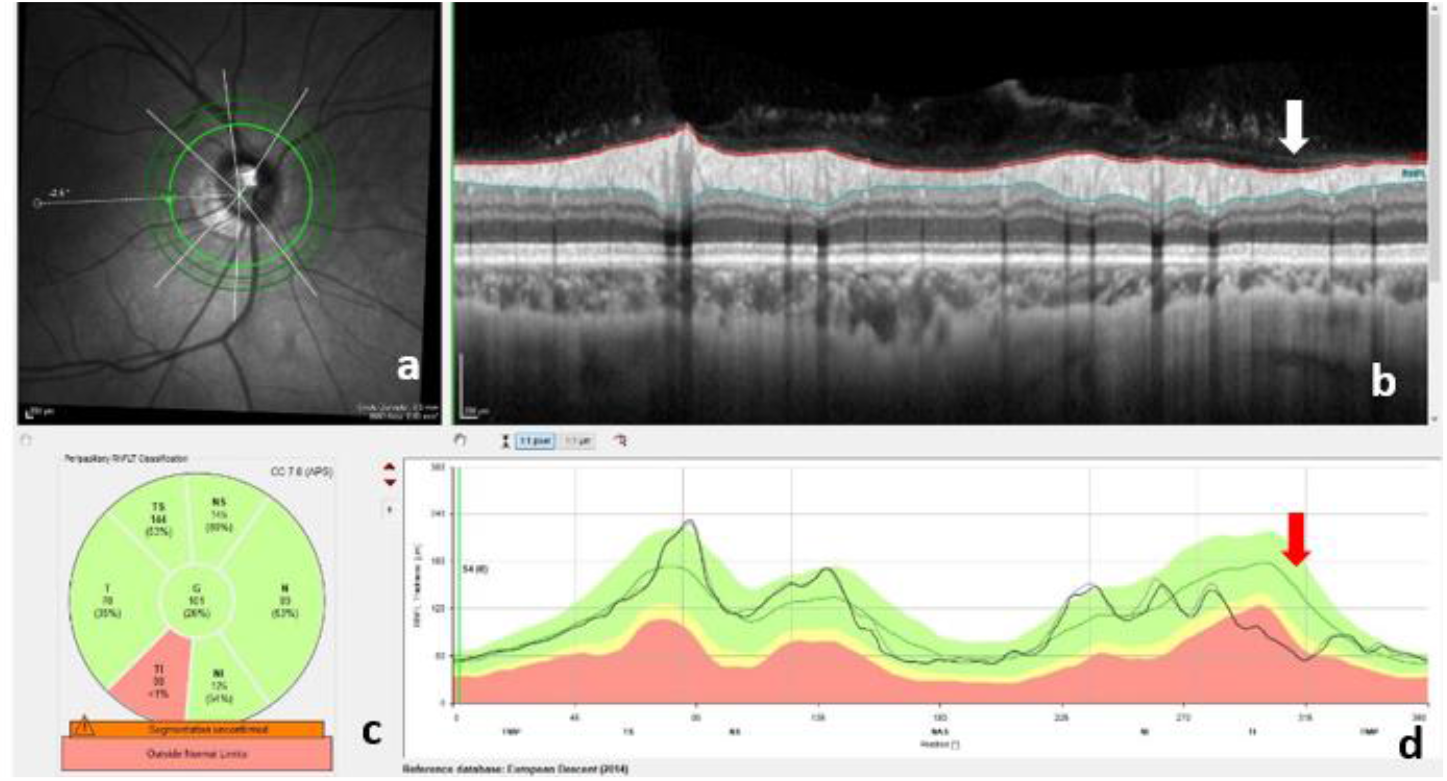
(a) Circumpapillary OCT scan in a clockwise direction from a 3.5 circle diameter of the center of ONH (b) Cross-sectional Spectralis TSNIT OCT B-scan (c) 6-sector Garway-Heath results (d) TSNIT profile; the red area of the 6-sector Garway-Heath indicates the circumpapillary RNFL is significantly thinned in the temporal-inferior sector and this defect is also visible on the B-scan and the TSNIT profile (white and red arrows).

Glaucomatous damage has a predilection for early damage in the inferior and superior zones.^7^ Glaucomatous loss of the RNFL leads to local defects, which are evident in the TSNIT OCT B-scan of Figure 1(b) (white arrow), 6-sector Garway-Heath analysis of Figure 1(c) and the TSNIT profile red arrow of Figure 1(d). Local thinning of the retinal layers in glaucoma varies in location, depth or width. With more advanced diseases, thinning becomes widespread, affecting all quadrants of the peripapillary area.^8^ Deep learning (DL) has shown promising performance in classifying 2D images to distinguish the glaucomatous ONH, such as an enlarged cup-to-disc ratio (CDR),^9^ which is deemed to be one of the crucial clinical features in glaucoma diagnosis and assessment. Initially, most studies have explored the DL segmentation to derive CDR from fundus photographs. Li and colleagues^10^ used the pre-trained Inception-v3 DL model to detect referable glaucomatous optic neuropathy and segment the cup and disc boundaries from fundus images. Similarly, Miri and co-workers^11^ used a multisurface graph-based technique to segment multimodal features. They trained the features using a random forest classifier to detect the optic disc and cup boundaries by combining both fundus and SD-OCT images. The limitation of these approaches is the known overlap between CDRs in normals and those with glaucoma.

OCT is now entrenched as an indispensable non-invasive imaging approach in the clinic. Cross-sectional OCT B-scans permit in vivo histopathological imaging of the retina and ONH and allow mapping of contours and topography.^12,13^ Different ONH features imaged using OCT, such as average retinal nerve fiber layer (RNFL) thickness, disc topography, ganglion cell layer, inner plexiform layer thickness, ganglion cell complex (GCC) layer thickness and macular RNFL thickness, are widely used in clinical features to train an artificial neural network in glaucoma detection.^14^ According to Christopher et al.^15^, unsupervised machine learning-based principal component analysis (PCA) could be a novel technique to detect glaucomatous nerve fibre layer damage using wide-angle OCT. Wang and colleagues^16^ developed a new BLNet (Boundary detection and Layer segmentation Network) model to detect all the retinal boundaries and segment all the layers from peripapillary circular optic nerve scans. It measured the RNFL thickness to classify glaucoma using a CNN and achieved an area under the receiver operating characteristic curve (AUC) of 0.864. Researchers at the National University of Singapore^17^ developed a custom U-net called DRUNET, a robust segmentation framework that can segment six ONH tissue layers by capturing both the tissue texture and spatial arrangement of tissues. The authors in another study^18^ developed a Branch Residual U-Network (BRU-Net), which combines residual building blocks with dilated convolutions into an asymmetric U-shape model which can successfully segment several retinal layers. This model reduced the training and validation mean square error (MSE) loss and improved the convergence speed compared to U-net.^19^

However, with all these methods, the accuracy of the model depends on the accurate segmentation and quantification of the relevant retinal structures of OCT images. The use of human-led image annotation potentially decreases classification accuracy. Machine-based segmentation also often failed due to the presence of speckle noise,^20-22^ low signal strength,^23^ motion artifacts,^24^ and vitreous opacity or vitreoretinal interface effects.^25-27^ Furthermore, mis-segmentation occurs in eyes with advanced glaucoma or retinal pathology (e.g., diabetic retinopathy).^28^

The benefit of using CNNs is their feature-agnostic ability, i.e., the ability to classify images by learning the position and scaling the variant structures within the data. This ability decreases the need for human input by eliminating manual segmentation and pre-processing. Recently, two studies reported that a feature agnostic CNN model performs better when processing unsegmented SD-OCT scans than segmented retinal layers^29^ to detect glaucoma. Maetschke and colleagues28 proposed a DL model that can detect glaucoma directly from raw OCT volumes of the ONH using a 3D CNN with an AUC of 0.94. The second study by Thompson and colleagues^30^ achieves a higher AUC (0.97) using the CNN model than the global RNFL thickness model (0.87) for automated glaucoma detection, trained from raw SD-OCT peripapillary B-scan images.

Deep learning models are often criticized as “black boxes” due to their failure to provide a reasonable explanation behind the prediction of a specific diagnosis. To address this gap between DL and clinician-based assessments, different visualization techniques such as Deconvolution Network (DeconvNet),^31^ Deep Visual Explanation (DVE),^32^ Local Interpretable Model-Agnostic Explanations (LIME),^33^ Class Activation Mapping (CAM),^34^ Gradient-weighted-CAM (Grad-CAM),^35^ etc. have been developed to explain and visualize the neural network mechanism and feature maps responsible for network prediction.

In this study, we aimed to explore the low-level and high-level learning features of different layers of CNN using pre-trained models to explore whether the raw TSNIT profile can distinguish glaucomatous eyes from healthy eyes. The models were fine-tuned to detect glaucoma from both raw and segmented (Inner Limiting Membrane (ILM)+RNFL layer) images using the transfer learning method. To the best of our knowledge, this is the first study using the ONH TSNIT retinal structural OCT feature for detecting glaucoma using deep learning and our preliminary work achieved promising result.^36^ The work has been substantially extended by implementing three different pre-trained models for i) glaucoma detection ii) CNN feature visualization and iii) localization of distinguishing retinal regions using the Grad-CAM technique by employing more datasets.

## 2. Materials and Methods

### Datasets

This study was a retrospective, a total of 1560 circumpapillary RNFL (cpRNFL) ONH OCT B-scans (Heidelberg Spectralis OCT; Heidelberg Engineering, Heidelberg, Germany), consisting of 780 segmented (ILM+RNFL) and 780 raw images of 370 glaucomatous and 410 normal eyes were collected. The patients were seen at the Centre for Eye Health, UNSW from 2015 to 2018, normal aged 49±13 and glaucoma aged 63±13 under the ‘Glaucoma Management Clinic’ (GMC). The glaucoma patients were diagnosed and managed by the GMC referred by the optometrist, CFEH optometrists and an ophthalmologist following clinical guidelines.^37^ A mixed dataset was selected with different types of glaucoma (e.g. primary open-angle glaucoma (POAG), normal-tension glaucoma (NTG), secondary open-angle glaucoma (SOAG), and angle-closure glaucoma (ACG) with different stages (early to severe) of glaucomatous optic neuropathy. The glaucoma staging was diagnosed by the clinicians, followed by the Mills criteria.^38^ The study adhered to all the tenets of the Declaration of Helsinki and was approved by the UNSW Sydney ethics committee. A written informed consent was obtained from each patient prior to using their de-identified clinical data in the study. The TSNIT retinal B-scan was cropped from the original machine’s cpRNFL OCT scans and resized using the bicubic interpolation method to desired pixels size using ImageJ software (https://imagej.net/Fiji): no other pre-processing was performed.

### Glaucoma detection

Pre-trained models were trained using 678 raw and segmented images (Normal: 364, Glaucoma: 314) for automated glaucoma detection. To balance the groups, 50 images from the glaucoma group were augmented by rotating 5 degrees clockwise. Finally, 728 images were split into 70% for training and 30% for validation. The remaining 102 images (46 normal and 56 glaucoma: early 31, advanced and moderate 23, severe 2) were used in the testing dataset to observe the performance of the model. The three pre-trained models SqueezeNet,^39^ ResNet18,^40^ and VGG16,^41^ were fine-tuned to train the images for feature visualization and glaucoma detection. The SqueezeNet model was selected for its simple architecture, reliance on few parameters, and fast learning abilities. The ResNet and VGG are powerful DL models and are widely used for medical image classification.^42^ The SqueezeNet model consists of 68 layers: only the last layer of the model is replaced with a new convolutional layer updating to two class numbers. The weight and biases of the fully connected layer are changed; all other remaining layers’ weights and biases are unchanged. The TSNIT images were resized to 227×227 pixels according to the input size of the SqueezeNet model. A minibatch size of 12, total epochs of 10, and a learning rate of 0.0002 were set for the final model.

The ResNet18 consists of 71 layers and is popular for its residual learning; gradients can flow through the shortened effective paths to any other earlier layer spontaneously, resolving the vanishing gradients problem for large and deep neural networks. For ResNet18, a minibatch size of 10, total epochs 10, and a learning rate of 0.00001 were selected for final classification. VGG16 consists of 41 layers and has demonstrated promising results for retinal OCT image analysis compared to other pre-trained models.^43-46^ For VGG16 the dataset was split into 80% training and 20% validation to avoid the overfitting problem. The batch size was 10, maximum epochs 10, learning rate 0.00005 were set to train the model. The AUC, calculated from the receiver operating characteristics (ROC) curve and confusion-matrix (CM) were used for performance evaluation. The classification metrics were calculated from the CM.

### Visualizing features map of TSNIT OCT

The technique of visualizing the learned features of CNN is termed feature visualization. We investigated how raw image features are learned throughout different layers of the network through the visualization of features from low-to high-level. The feature maps were visualized through the different layers of the SqueezeNet model (Figure 2). To visualize the features through the neural network’s convolutional filters and activations maps, SqueezeNet was preferred because it has fewer parameters, a low run-time facility with limited memory. Both glaucomatous and healthy TSNIT raw and segmented OCT images were used to feed the network and observe the activations of different layers of the network. The CNN features were investigated by observing the activated image regions in the convolutional layers and compared to the corresponding regions of the raw images. The activation output was normalized using the ‘mat2gray’ function (convert the matrix into grayscale images) and the minimum activation value was set between 0 to 1 to provide a clear visual explanation.

**Figure 2.**
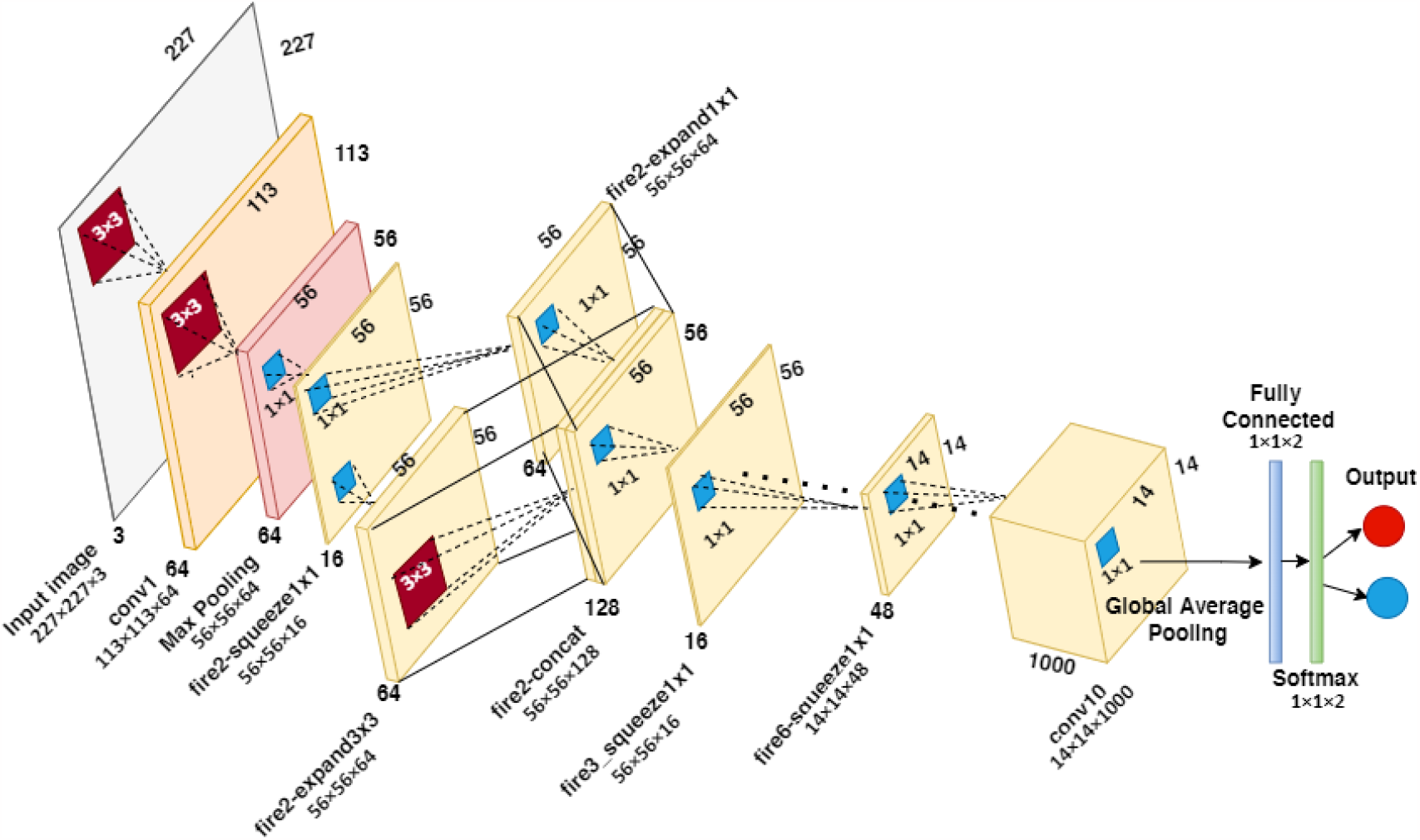
SqueezeNet model diagram for feature map visualization and glaucoma detection.

The first convolution layer and two deeper layers of the CNN were selected and demonstrated the features of the strong activation channel for both normal and glaucomatous eyes images. The first convolutional layer consists of different convolution filters (kernel size: 3×3) and performs image operations such as image blurring, sharpening, embossing, edge detection, etc., to learn the useful image elements. The network often learns one feature per channel, or filter. For SqueezeNet, the first convolutional layer has 64 channels (113×113×64) (Figure 2). We first examine the output activations of the first convolutional (conv1) layer on an 8×8 grid, to display 64 images (one for each channel of the layer: see Figure 3). Among the 64 channels, the strongest activation channel is found by calculating the maximum pixel values of the conv1 layer and displaying it with the same size as the original image (Figure 3 right).

**Figure 3.**
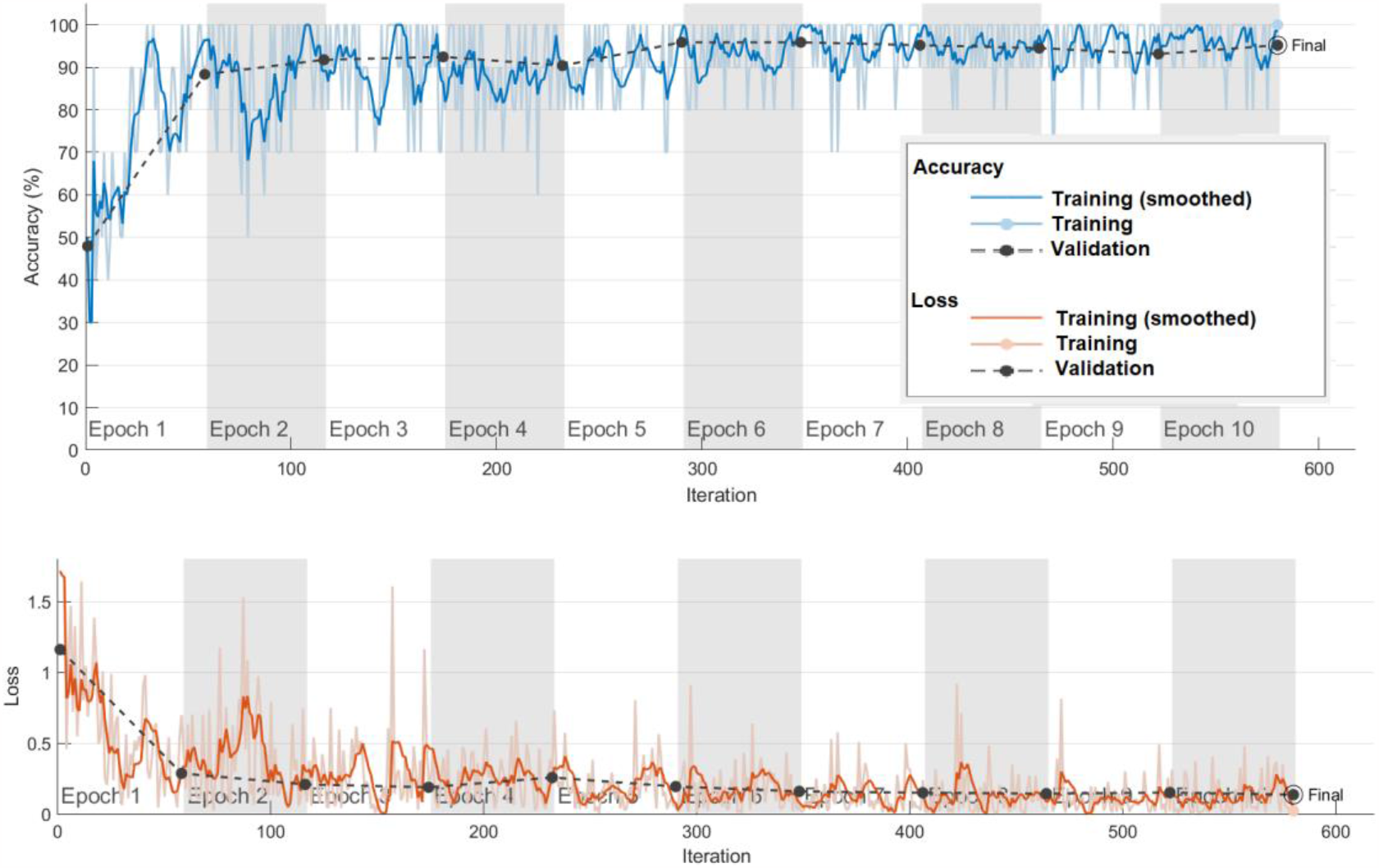
The accuracy and loss curve during the training progress of the VGG16 model.

### Investigation of network predictions using Grad-CAM

The pre-trained models trained using both segmented and raw OCT images were investigated with Grad-CAM activation function to visualize which regions of the images are counted for prediction of the two classes. Grad-CAM creates a class-specific localization map highlighting the features of the image using the gradient of the prediction score of the specific class with respect to the last convolutional layer of the model. The Grad-CAM produces heatmap transparently on the image by using ‘AlphaData’ and ‘jet’ colormap to generate deep red as the peak value (greatest impact on the classification) and deep blue as the lowest class value.

## 3. Results and Discussion

### Glaucoma detection

The classification results of the validation and test images of the three fine-tuned pre-trained models are demonstrated using the CM in Table 1 and performance metrics in Table 2. The AUC was calculated from the ROC curve comparing the true positive rate to the false positive rate. The sensitivity, specificity, precision, and F1-score were calculated from the CM based on four outcomes: true positive-(correctly identified as glaucoma); true negative- (correctly identified as normal); false positive- (normal incorrectly classified as glaucoma); false negative- (glaucoma incorrectly classified as normal). The segmentation of ILM+RNFL has no impact on the results of DL models. Therefore, we are only demonstrating the results of raw OCT images.

**Table 1.**
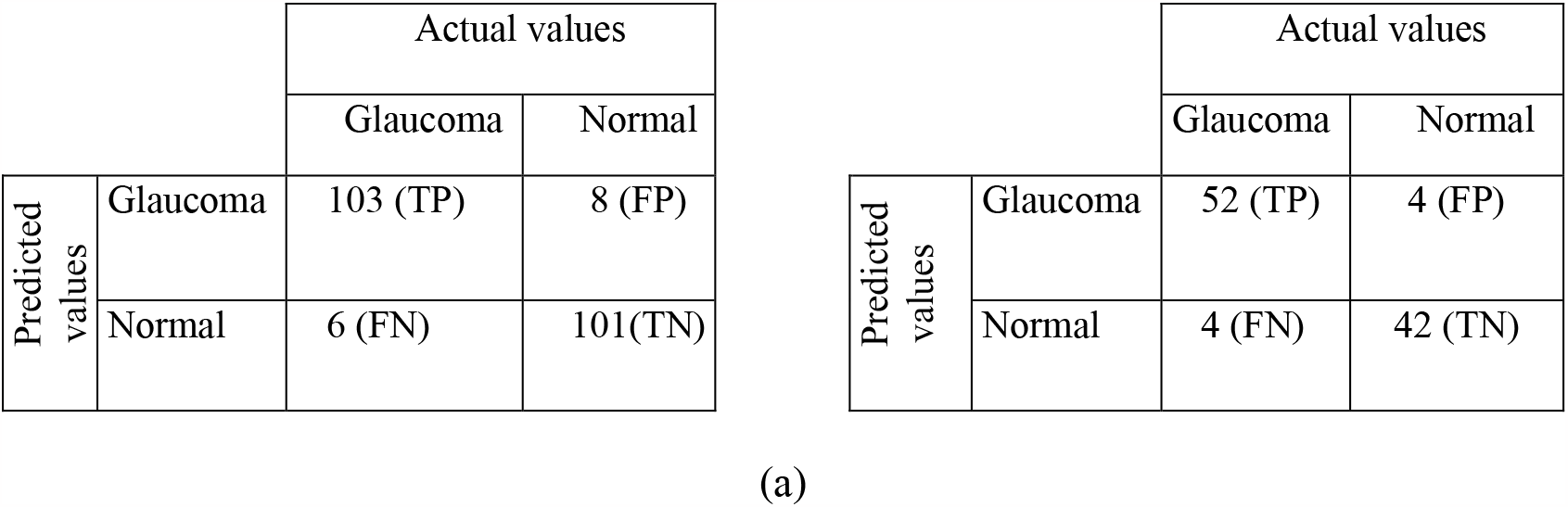
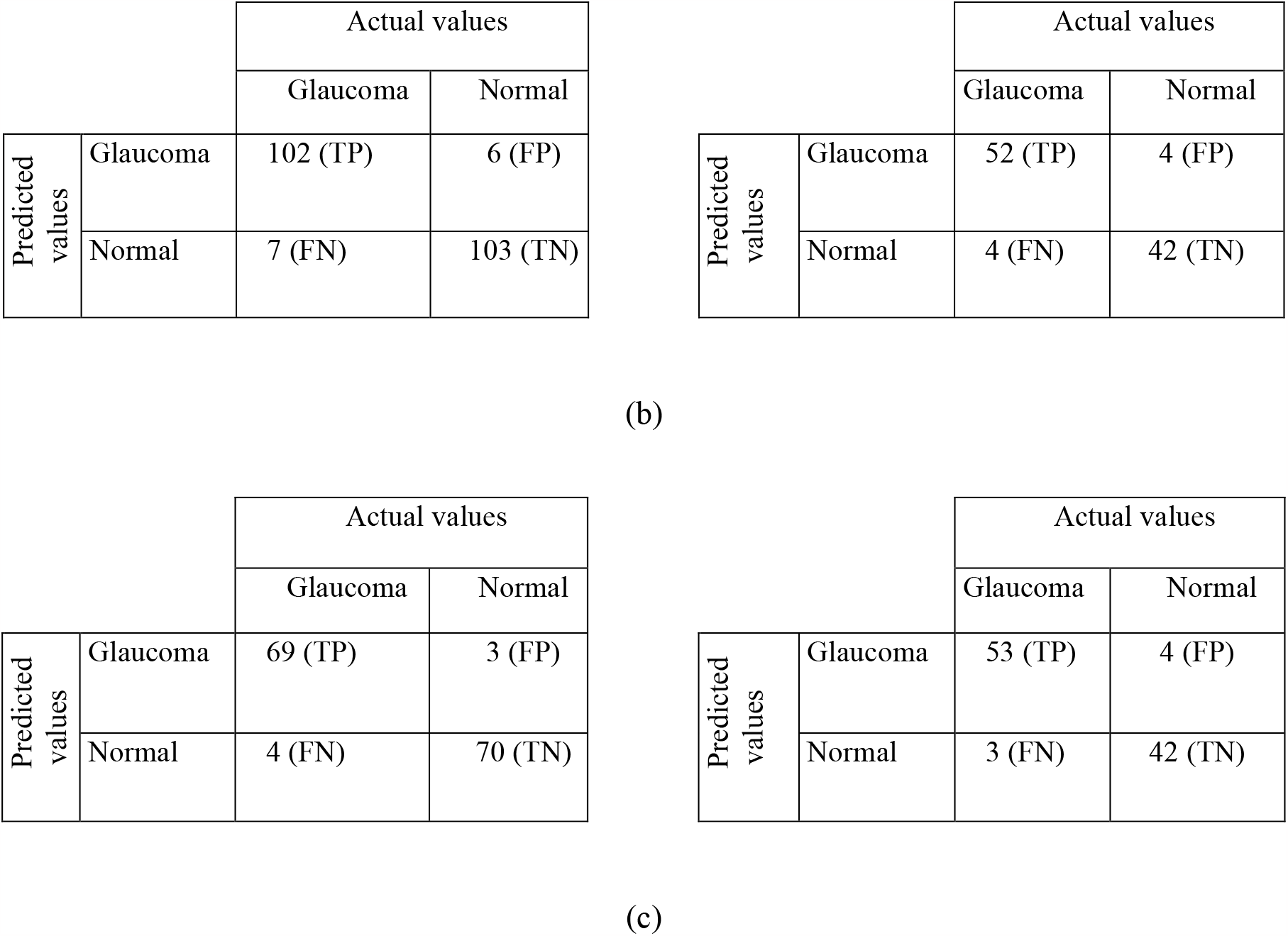
Confusion matrix of validation data (left) and test data (right) for three models (a) SqueezeNet (b) ResNet-18 (c) VGG16.

**Table 2.** Comparison of pre-trained models’ performance for glaucoma detection using TSNIT OCT images

| CNN | AUC | Accuracy (%) | Sensitivity (%) | Specificity (%) | Precision | F1-score |
| --- | --- | --- | --- | --- | --- | --- |
| SqueezeNet | 0.973 | 93.6% | 94.5% | 92.7% | 92.8% | 93.6% |
| ResNet-18 | 0.978 | 94.0% | 93.6% | 94.5% | 94.4% | 94.0% |
| VGG16 | 0.988 | 95.2% | 94.5% | 95.9% | 95.8% | 95.0% |

Figure 3 demonstrated the accuracy and loss curve of the VGG16 model against each epoch, which demonstrated that the model was robustly trained and obtained a stable validation accuracy. The CMs (Table 1), Table 2 and the ROC curve of Figure 4 demonstrate that of the pre-trained models, VGG16 offered the best performance, with the highest AUC (0.98), accuracy (95.2%), sensitivity (94.5%), specificity (95.9%), precision (95.8%) and F1-score (95.0%) on validation data. The accuracy for test data was 93.0%, sensitivity 94.6%, specificity 91.3%, precision 93.0% and F1-score 93.8%. Among 102 images, the VGG16 model correctly identified 53 glaucomas out of 56 images; only three images from eyes with early glaucoma were misclassified as normal. Further image assessment found the 6-sector Garway-Heath results also look normal for two glaucoma eyes, no affected region was found for these two earlier glaucomatous eyes. Four images from normals were incorrectly classified as glaucoma. Of these, one eye was suspected of macular degeneration in the clinical record, two had a family history of glaucoma, and for the other one had no ocular or family history of note. Most of the test results included early glaucoma (31 out of 56); therefore, we can say the DL model using TSNIT OCT images may detect early glaucoma could therefore help clinicians initiate earlier treatment.

**Figure 4.**
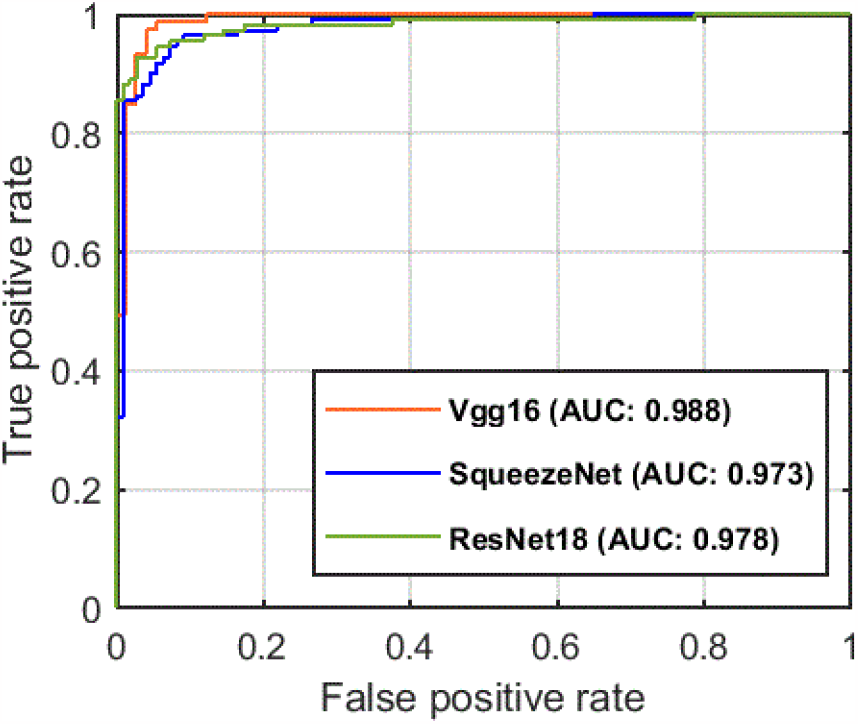
The ROC curve for three transfer-learning models for automated glaucoma detection.

### Feature map visualization

Figure 5 demonstrates that the strong activation channel appears similar for raw and segmented images. However, in the conv1 layer few channels are only activated with segmented edges. The features of the conv1 layer will pass through several rectified linear units (ReLu), max-pooling and convolution filters throughout the network. The Relu only activates on positive pixel values and assigns zero for negative feature map pixel values.^47^ The max-pooling function reduces the feature map sizes by calculating the maximum pixel value within its stride window^48^ whilst also producing the most important features of the images in the network. Therefore, the most significant features are activated and pass to the adjacent layers. Figure 6 examines the deeper layer activations of the SqueezeNet model. Among 68 layers, we chose the 35 number layer (fire6-squeeze1×1) - which is a convolutional layer - and the last convolutional layer (relu_conv10) to observe which features were finally used by the network to make the classification decision. For the fire6-squeeze1×1 layer, images are displayed on a 6×8 grid (one for each channel of the layer). The features map (14×14×48) of this layer was ambiguous and combined features of the previous layers. For relu_conv10, only the strong activation channel is displayed in Figure 6 (right).

**Figure 5.**
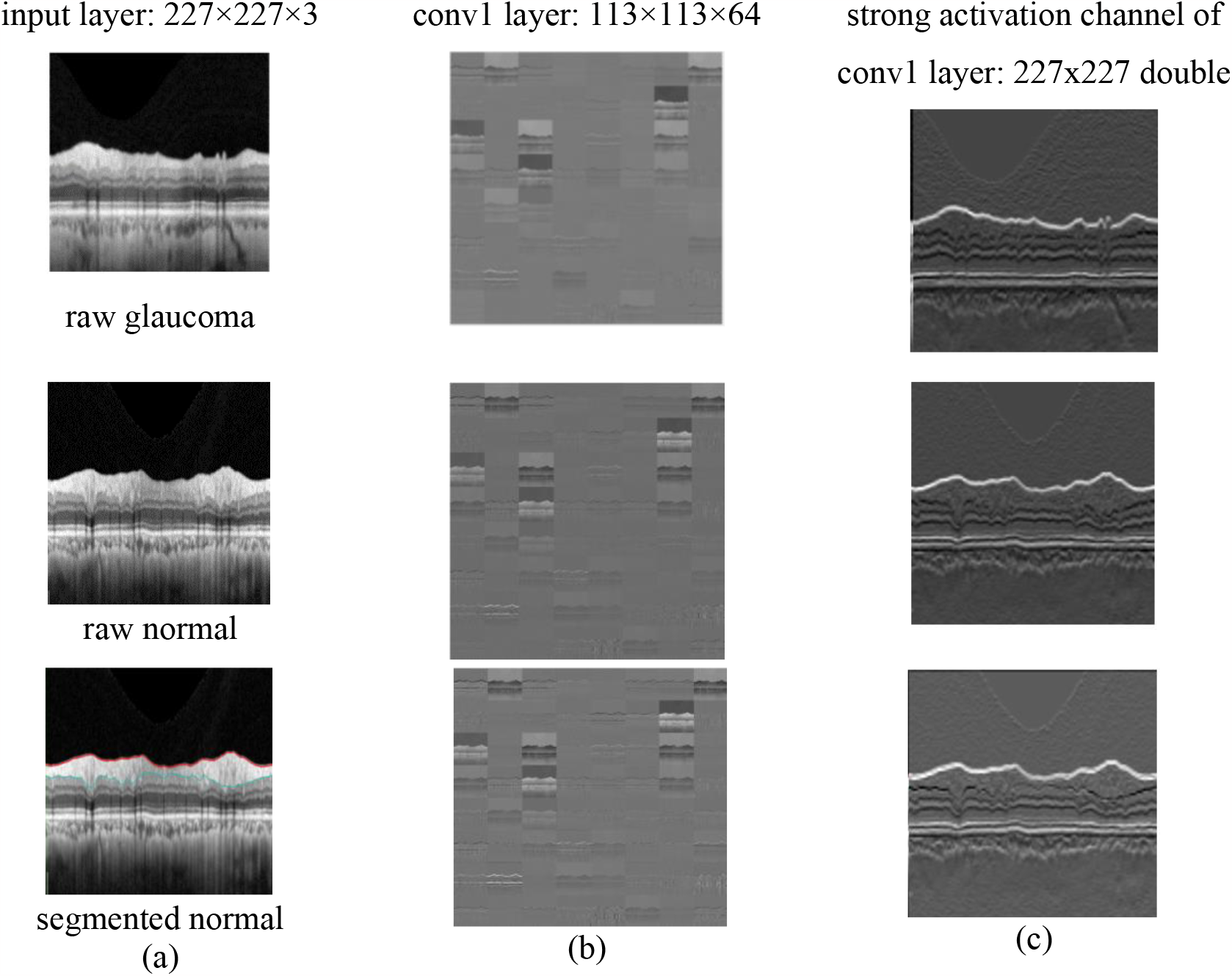
a) Original OCT images for normal and glaucomatous eyes (b) The activation channels of the first convolutional layer (c) The strongest activation channel of first convolutional layer.

**Figure 6.**
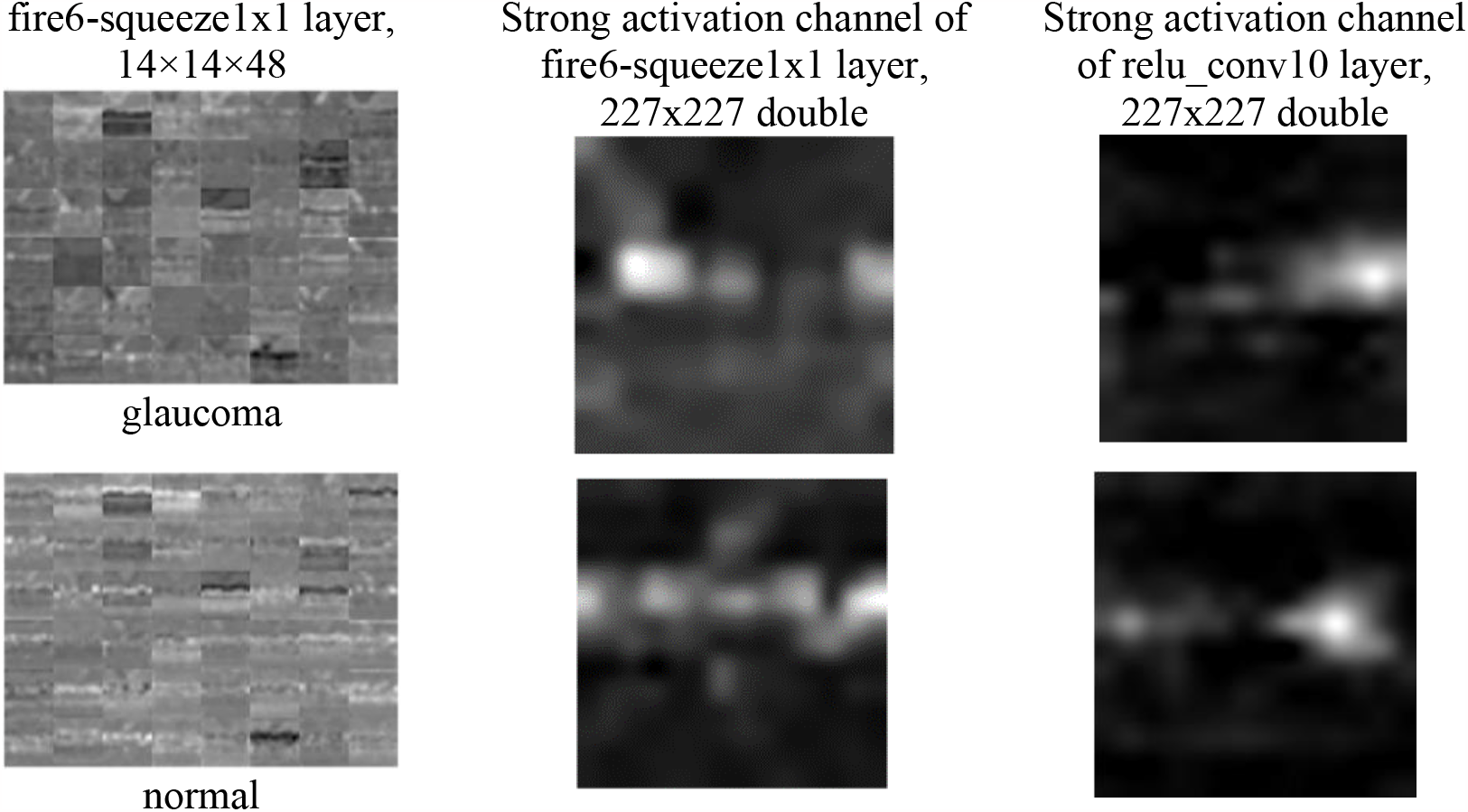
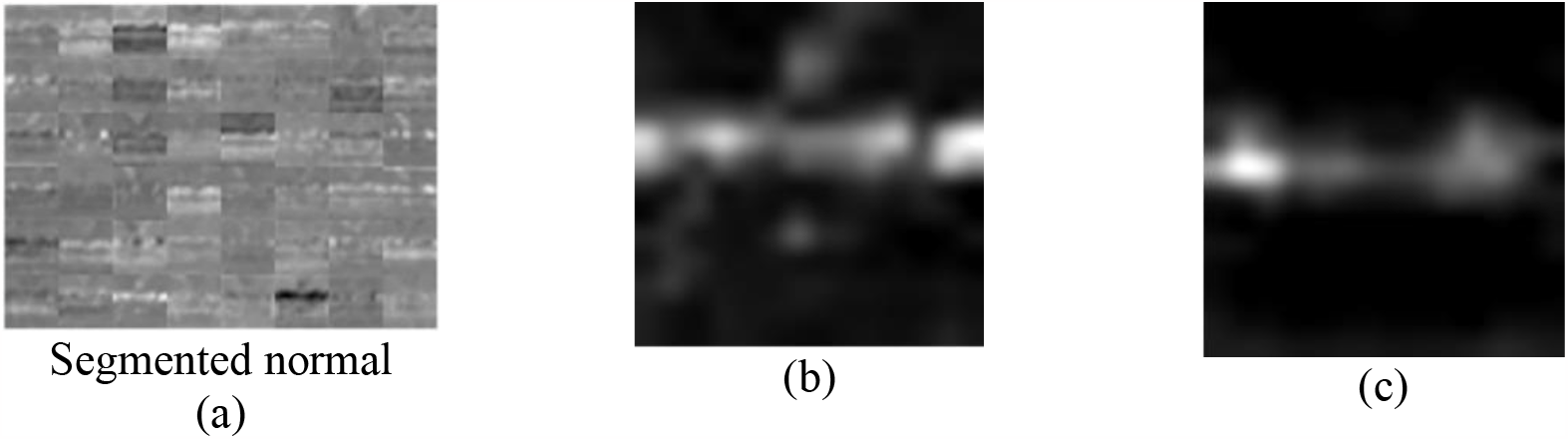
(a) The activation channels of fire6-in squeeze1×1 convolutional layer (b) The strongest activation channel of fire6-in squeeze1×1 convolutional layer (c) The strongest activation channel of the last convolutional layer.

If we compare the original image with the strong activation channels (Figure 6), we can see the channel activates most of the RNFL layer of the image. The active regions of positive (white pixels) are bigger in the normal image’s activation channel than in the glaucomatous image’s. This shows that CNN correctly locates the affected region (glaucoma-the brightest pixels) and the healthy RNFL regions in normal eyes images (both raw and segmented) in the final convolution layer (conv10).

### Investigate network predictions using Gradient-weighted-Class Activation Mapping (Grad-CAM)

We explored the features of the images which were considered by the three models when predicting the two groups. Four random glaucomatous and normal images were investigated using Grad-CAMs, and the results are demonstrated in Figure 7 and Figure 8.

**Figure 7.**
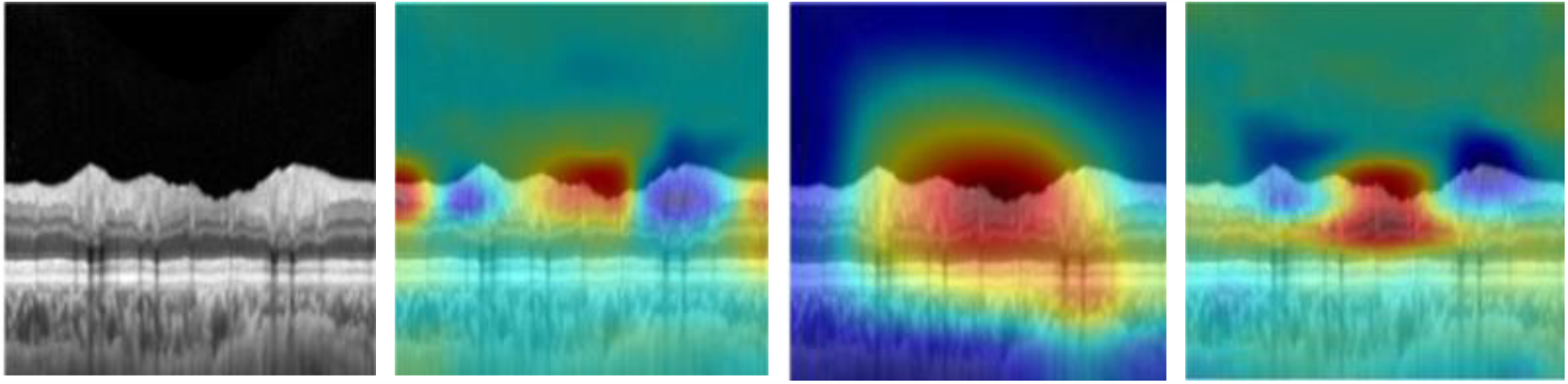
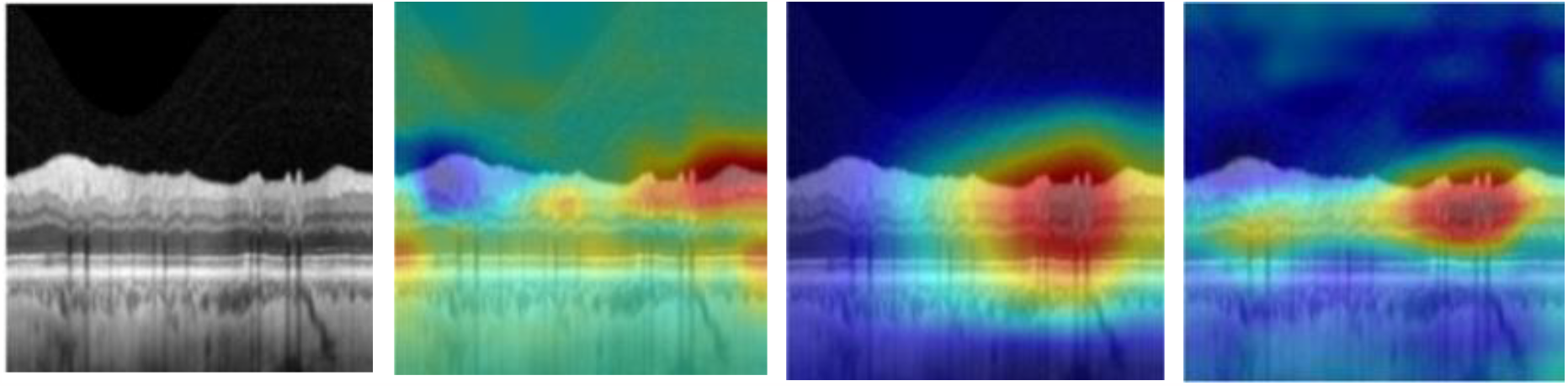
The Grad-CAM result for SqueezeNet, ResNet18, and VGG16 (left to right) extracted from raw images of glaucomatous eyes (left).

**Figure 8.**
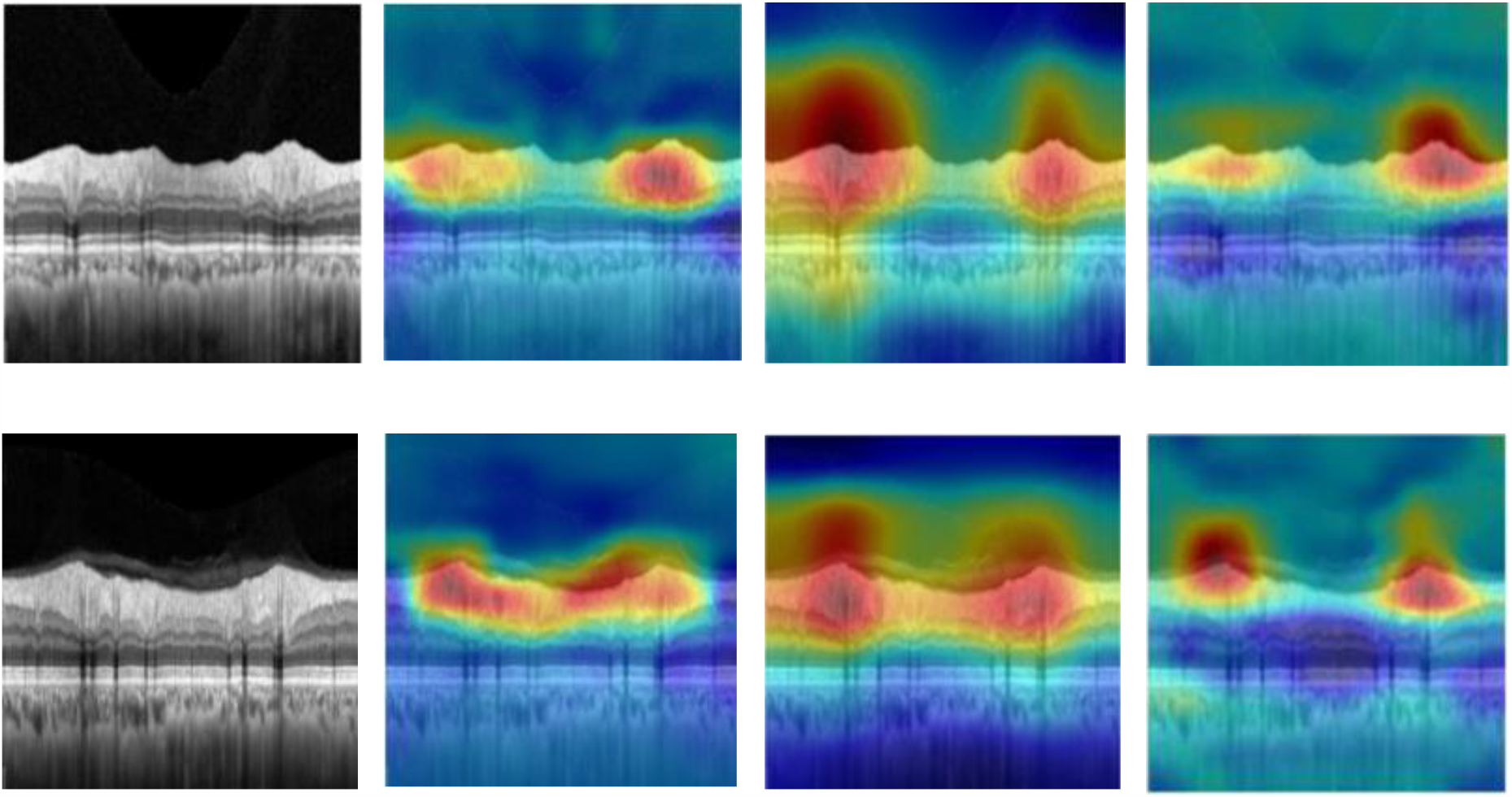
The Grad-CAM result for SqueezeNet, ResNet18, and VGG16 (left to right) extracted from raw images of normal eyes (left).

In Figure 7 and Figure 8, we can see that the heatmaps generated from the Grad-CAM accurately differentiate the two groups. The localized region for glaucoma is in line with the definition of glaucoma. Thinning of the RNFL indicates glaucomatous damage, whilst a thicker RNFL suggests is present in normals. The three models precisely identify the glaucomatous damage across the TSNIT scan. We also explored Grad-CAMs for segmented images, and the heatmaps were similar for both raw and segmented images. Thus, we conclude that the raw TSNIT OCT-based DL model can contribute to glaucoma diagnosis. The CNN detects the same structural information as the expert clinicians without consideration of prior clinical information.

This study explored automated glaucoma detection using three pre-trained models, of these, VGG16 showed the most promise, with an AUC 0.98 for validation data. The sensitivity was 95% in the test dataset, and the F1 score was 94%, which is better than that found in previous studies.^26,28,45^ Visualization of image features from low to high levels demonstrated that the RNFL layer was the most discriminative feature among all the retinal layers. To the best of our knowledge, this is the first study where the CNN features of retinal OCT images were visualized and investigated. The Grad-CAM reveals that the three models successfully localized the regions affected by glaucomatous optic neuropathy (red regions are highly predictive for specific classes) and can separate glaucomatous from normal eyes, which is consistent with previous literature.^45^ The OCT TSNIT scan is ideal for glaucoma imaging as it encompasses the complete retinal structure throughout 360 degrees in the peripapiallary region. Our results demonstrate that the raw version of this scan can be used to differentiate early glaucoma using deep learning techniques. Moreover, our results suggest that DL models can detect glaucoma via a feature agnostic approach from TSNIT OCT B-scans. Hence, these DL algorithms do not require any machine or human-led segmentation or pre-processing before training, which may be an attractive feature in large scale screening applications. Use of a single cpRNFL OCT scan could streamline glaucoma screening, improve the experience for patients and better optimize health resources.

We note that our study has limitations. Our study is a single-centre study; most of the patients in this study are Caucasian and Asian; therefore, we could not investigate the effect of ethnicity on the retinal layer. Moreover, we did not examine the glaucoma TSNIT layer changes with other concomitant diseases (like cataracts, diabetes etc.). Therefore, there is a need for additional, global validation of deep learning models for different ethnic backgrounds including larger age groups and further investigation regarding concurrent ocular diseases’ impact on TSNIT scans. Furthermore, the robustness of the DL model using TSNIT OCT scan with improved results could be achievable in the future by integrating larger training datasets.

## 4. Conclusion

In conclusion, the classification results and the interpretation of optical coherence tomography images using pre-trained deep learning models demonstrated promising and reliable performance superior to comparable studies in this field. This suggests that the Temporal-Superior-Nasal-Inferior-Temporal retinal profile could be considered a novel clinical imaging feature to train artificial neural networks for automated glaucoma detection and management. The features visualization and localization process solved the ‘black box’ problem of artificial intelligence and renders the classification process more transparent to users. Interpretable results can help shed new perspectives to clinicians during the diagnostic phase and increase the reliability of the deep learning model at the clinician’s level. This automated transparent deep learning model using Temporal-Superior-Nasal-Inferior-Temporal retinal optical coherence tomography images could be a powerful tool that may ultimately improve screening for glaucoma, even in its early stages.

## Data Availability

All data produced in the present study are available upon reasonable request to the authors

## Acknowledgements

We would like to thank CFEH, UNSW, Sydney for assisting in the data collection. Clinical services at the CFEH are funded by Guide Dogs NSW/ACT. Guide Dogs NSW/ACT had no role in study design, data collection and analysis, decision to publish, or manuscript preparation.

